# Data independent acquisition mass spectrometry (DIA-MS) analysis of FFPE rectal cancer samples offers in depth proteomics characterization of response to neoadjuvant chemoradiotherapy

**DOI:** 10.1101/2023.05.12.23289671

**Authors:** Aleksandra Stanojevic, Martina Samiotaki, Vasiliki Lygirou, Mladen Marinkovic, Vladimir Nikolic, Suzana Stojanovic-Rundic, Radmila Jankovic, Antonia Vlahou, George Panayotou, Remond J.A. Fijneman, Sergi Castellvi-Bel, Jerome Zoidakis, Milena Cavic

**Affiliations:** Department of Experimental Oncology, Institute for Oncology and Radiology of Serbia, Belgrade, Serbia; Institute for Bioinnovation, Biomedical Sciences Research Center “Alexander Fleming”, Vari, Attica, Greece; Department of Biotechnology, Biomedical Research Foundation, Academy of Athens, Athens, Greece; Clinic for Radiation Oncology and Diagnostics, Department of Radiation Oncology, Institute for Oncology and Radiology of Serbia, Belgrade, Serbia; Clinic for Medical Oncology, Institute for Oncology and Radiology of Serbia, Belgrade, Serbia; Faculty of Medicine, University of Belgrade, Serbia; Department of Pathology, The Netherlands Cancer Institute, Amsterdam, the Netherlands; Gastroenterology Department, Fundació Clínic per la Recerca Biomèdica-Institut d’Investigacions Biomèdiques August Pi i Sunyer (FRCB-IDIBAPS), Centro de Investigación Biomédica en Red de Enfermedades Hepáticas y Digestivas (CIBERehd), Hospital Clínic, University of Barcelona, Barcelona, Spain; Department of Biology, National and Kapodistrian University of Athens, Greece

**Keywords:** data-independent acquisition mass spectrometry, neoadjuvant chemoradiotherapy, rectal cancer, proteomics

## Abstract

**Background:** Understanding the molecular features associated with response to neoadjuvant chemoradiotherapy is an unmet clinical need in locally advanced rectal cancer (LARC). The aim of the study was to apply a high-sensitivity proteomic approach for in-depth characterization of the LARC proteome in search of patients who might have a good response to preoperative treatment and potentially be followed by a watch-and-wait strategy, rather than having immediate surgery, maximizing the therapeutic effect and quality of life.

**Methods:** A total of 97 LARC patients treated at the Institute for Oncology and Radiology of Serbia in the period of 2018-2019 were included in the study. Patients were treated with long-course chemoradiotherapy (CRT): Radiotherapy (RT) was delivered with a total dose of 50.4 Gy in 28 fractions; concomitant chemotherapy (5-FU, 350 mg/m^2^ daily) and Leucovorin (25 mg/m^2^ daily) was administered during the first and the fifth week of RT. Patients were evaluated in week 6-8 after treatment completion with pelvic MRI scan and rigid proctoscopy. Pathohistological response after surgery was assessed according to tumor regression grading (TRG) categories by Mandard. Twenty biopsy samples taken at diagnosis were used for proteomic analysis, 9 responders (R, TRG 1-2), and 11 non-responders (NR, TRG 3-5), to achieve the maximum range of different molecular features potentially associated with response. Formalin-fixed paraffin-embedded (FFPE) biopsies were processed, and isolated proteins were digested with trypsin. The resulting peptides were analyzed by liquid chromatography coupled to a Q Exactive HF-X mass spectrometer operated in data independent mode (DIA-MS). Data analysis was performed with DIA-NN and Perseus. Data are available via ProteomeXchange with the identifier PXD040451.

**Results:** The use of DIA-MS allowed the identification and quantification of more than 3,000 proteins per sample in general, a significant increase when compared to the 1,000 proteins previously identified by Data Dependent Acquisition-MS (DDA-MS) in LARC FFPE samples. In total, 4,849 proteins were identified in 20 rectal cancer FFPE samples. Principal Component Analysis (PCA) indicated that responders had a significantly different proteomic profile than non-responders. Statistical analysis of the two groups resulted in the identification of 915 differentially expressed proteins (DEPs) (215 in responders and 700 in non-responders, p<0.05), and 384 with more stringent criteria (p<0.01). Results indicate that some of the leading signaling pathways that correlate with response include the metabolism of RNA, MYC targets, neutrophil degranulation, cellular transport, and response to stimuli.

**Conclusions:** The DIA-MS approach offered unprecedented proteome coverage for FFPE samples. The differentially expressed proteins and biological processes constitute interesting findings that hold the potential for improving LARC patient management.

## Introduction

Colorectal cancer (CRC) is the third most common type of cancer worldwide with almost two million newly diagnosed cases in 2020(1). High mortality rates place CRC in second place after lung cancer (1). In Serbia, the situation is similar. According to data from the Institute of Public Health “Milan Jovanovic Batut” of the Republic of Serbia, CRC holds the second place by incidence and mortality rates, with around 5000 new cases and 3.000 deaths annually (2). In majority of cases, it is diagnosed in advanced stages where limited treatment options are available and survival is poor. Our group and others have invested efforts into profiling the diagnostic, prognostic and predictive factors for CRC and anal cancer, in an effort to provide better research strategies for treatment and overall management (3–7). However, current early detection and screening programs, as well as treatment options, need further improvement on a global level. Rectum is the most distal part of the digestive tract located between the sigmoid colon and anal canal. Colon and rectal cancers have been traditionally considered as a single disease entity and rectal cancer (RC) represents around 35% of diagnosed CRC cases. Rectal cancer has distinct environmental and genetic risk factors that differentiate it from colon cancer (8). Its incidence has been reported to increase in the 18 to 50-year age group, especially in younger adults (9).

Locally advanced rectal cancer (LARC) is the most diagnosed type of RC, which includes stage II (T3/4N0M0) and III (T1-4N+M0) according to the Union for International Cancer Control (UICC)(10). The standard treatment for LARC is neoadjuvant chemoradiotherapy (nCRT) followed by radical surgery (total mesorectal excision). nCRT was established as the gold standard of LARC treatment after 2004 as a result of two studies CAO/ARO/AIO-94 and EORTC 22921, which compared it to previously used adjuvant radiotherapy with or without chemotherapy. According to the CAO/ARO/AIO-94, neoadjuvant RT dramatically reduced the rates of local failure, while the 11-year follow-up update showed that the long-term overall survival rate was about 60% (11)The EORTC 22921 trial showed that the use of chemotherapy with neoadjuvant radiation reduced local recurrence rates but had no effect on distant progression-free survival. Additionally, nCRT contributed to sphincter preservation and improved the patient’s quality of life (12). However, only 20%-30% of patients experience a complete clinical or pathological response to nCRT, while some patients will experience poor response or will have distant progression during nCRT (13,14). Characterization of mechanisms of response to therapy and the search for predictive biomarkers to nCRT is an unmet need in LARC.

The watch and wait approach was introduced because of the need for close follow-up of LARC patients with complete clinical response to nCRT, and allowed the extension of periods between neoadjuvant therapy and surgery, thus lowering morbidity related to surgery (15–18). No biomarker has yet been validated in this setting. We aimed to perform in-depth proteomics characterization of preoperative LARC biopsy samples by employing data-independent acquisition mass spectrometry (DIA-MS) to unravel new tissue molecular features that might lead to different responses to nCRT. Patients with a favorable response to nCRT would be candidates for a less invasive surgical approach or would be enrolled in a watch and wait approach in the case of a complete clinical response (cCR). That would increase their quality of life and contribute to the overall reduction of treatment costs (18).

## Experimental Design and Statistical Rationale

### Patient cohort characteristics and treatment

A total of 97 LARC patients treated at the Institute for Oncology and Radiology of Serbia in the period of 2018-2019 were included in the study. The inclusion criteria were histopathologically verified adenocarcinoma of the rectum, with a distant margin up to 12 cm from the anal verge by rigid proctoscopy, ECOG performance status ≤ 2. LARC was defined as T3-T4N0 or any T stage N+, according to clinical and histological criteria of the 8th edition of the TNM classification of malignant tumors (19). Pretreatment evaluation included an abdominal and pelvic MRI scan and a computed tomography (CT) scan or X-ray of the chest. All patients were treated with long-course chemoradiotherapy (CRT). Radiotherapy (RT) was delivered with a total dose of 50.4 Gy in 28 fractions (conventionally fractioned 1.8Gy/fr), using the technique with 3 or 4 radiation areas (all areas as recommended by the International Committee of Radiation Units and Measurements (ICRU, 50/62 per day) (20) Concomitant chemotherapy was initiated on the first day of RT and administered during the first and fifth week of RT. The chemotherapy regimen included: 5-FU (350 mg/m^2^, daily) and Leucovorin (25 mg/m^2^, daily). A complete patient medical database has been prepared from official records.

Patients were assessed for tumor response between the 6^th^ and 8^th^ week after CRT completion with pelvic MRI scan and rigid proctoscopy. The pathohistological response after surgery was assessed according to tumor regression grading (TRG) categories by Mandard (20). According to the TRG status, the patients were divided into two groups: responders (TRG 1-2) and non-responders (TRG 3-5). Our study included analysis of extreme candidates in order to achieve the maximum range of different molecular features potentially associated with response. Twenty-four formalin-fixed paraffin-embedded (FFPE) biopsy samples were taken at the moment of disease diagnosis and were collected and used for proteomic analysis. After the quality control check, 4 samples were excluded from further analysis, and finally, twenty samples were processed (9 responders and 11 non-responders). Characteristics of the study cohort are shown in Table 1.

**Table 1.** Clinical data of the study cohort.

| Characteristics |  | Responders<br>N (%) | Non-responders<br>N (%) | P-value* |
| --- | --- | --- | --- | --- |
| Gender |  |  |  |  |
|  | Male | 3 (33.3) | 7 (63.6) | 0.4 |
|  | Female | 6 (66.7) | 4 (36.4) |  |
| Age (years) |  |  |  |  |
|  | Mean (SD) | 64.0 (6.7) | 62.4 (10.3) | 1.0 |
|  | Median (Range) | 66.0 (50-72) | 64.0 (48-83) |  |
| UICC staging | II | 0.0 (0) | 2.0 (18.2) | 0.5 |
|  | III | 9.0 (100) | 9.0 (81.8) |  |
| Tumor grade |  |  |  |  |
|  | 1 | 7.0 (77.8) | 8.0 (72.7) | 0.8 |
|  | 2 | 2.0 (22.2) | 3.0 (27.3) |  |
|  | 3 | 0.0 (0.0) | 0.0 (0.0) |  |
| Tumor localization | Inferior rectum (<5cm) | 5.0 (55.6) | 9.0 (81.8) | 0.4 |
|  | Mid rectum (5-10cm) | 4.0 (44.4) | 2.0 (18.2) |  |
|  | Superior rectum (>10cm) | 0.0 (0) | 0.0 (0) |  |
| Acute toxicity |  |  |  |  |
|  | Without | 1.0 (12.5) | 2.0 (18.2) | 0.7 |
|  | With | 8.0 (87.5) | 9.0 (81.8) |  |
| Tumor Regression |  |  |  |  |
| Grade (TRG) |  |  |  |  |
|  | 1 | 8.0 (88.89) | / |  |
|  | 2 | 1.0 (11.11) | / |  |
|  | 3 | / | 2.0 (18.18) |  |
|  | 4 | / | 7.0 (63.64) |  |
|  | 5 | / | 2.0 (18.18) |  |
| Total |  | 9 (100) | 11 (100) |  |

\*Chi squared test with Yates correction (two-tailed).

The project was approved by the Ethics Committee of the Institute for Oncology and Radiology of Serbia (approval No. 2211-01 from 11.06.2020.) and all patients signed informed consent.

### Protein extraction from FFPE tissue samples

From each FFPE sample, 10 sections 15-20 μm thick were cut using a microtome and transferred to 2 mL Eppendorf tubes. From all sections, 3 were selected that contained the largest amount of tissue. Samples were deparaffinized using Xylene in two steps of 5 min and 1 min successively. The samples were centrifuged and in the next step, the pellet tissue was rehydrated using several dilutions of ethanol and finally washed with double distilled water. The tissue pellet was air-dried and dissolved in FASP protein extraction buffer (100mM Tris-HCl pH 7.6, SDS 4%, 100mM 1,4-Dithioerythtriol (DTE)). The tissue was homogenized, while the disintegration of the cell membrane was achieved by sonication (three cycles of 5 sec with 36% power). The samples were heated at 90℃ for 1 h to extract the cellular proteins into the solution. The supernatant containing extracted proteins was transferred to a new Eppendorf tube and an appropriate amount of ammonium bicarbonate (ABC) buffer was added. The sample was concentrated using a 3 kDa cut-off Amicon filter.

### Protein digestion and preparation for LC-MS/MS Analysis

The total volume of concentrated proteins was added on SDS PAGE (5% stacking gel, 12% separating gel). Preparative SDS PAGE was performed, and the gel was fixed, washed, and stained with Coomassie colloidal dye (Supp. Material 1). Protein bands were cut from the gel for each sample separately, chopped, and transferred to Eppendorf tubes. The strips were decolorized with a solution of 40% Acetonitrile, and 50 mM NH_4_HCO_3_ until parts of the gel became completely transparent. The samples were reduced with 10mM DTE in 100mM NH_4_HCO_3_ and alkylated 10mg/mL Iodoacetamide in 100mM NH_4_HCO_3_ and then washed with 100mM NH_4_HCO_3_, destain solution, and water, respectively. The samples were dried in a speed vac until transparent crystals formed. Each sample was treated with trypsin solution which enables the cutting of the polypeptides after the amino acids lysine and arginine. The peptides formed were extracted with NH_4_HCO_3_ solution followed by incubation in a 1: 1 solution of 10% Formic acid and Acetonitrile. The peptide solution was purified using PVDF filters (Merck Millipore). The samples were dried in a Speedvac and prepared for further processing.

### LC-MS/MS Analysis

Samples were run in two technical replicates on a liquid chromatography-tandem mass spectrometry (LC-MS/MS) setup consisting of an Ultimate 3000 RSLC online with a Thermo Q Exactive HF-X Orbitrap mass spectrometer. Peptide solutions were directly injected and separated on a 25 cm-long analytical C18 column (PepSep, 1.9 μm beads, 75 µm ID) using 90 minutes long runs using a gradient of 7% Buffer B (0.1% Formic acid in 80% Acetonitrile) to 36% was used for 70 min, followed by an increase to 95% in 5 min, and a second increase to 99% in 0.5 min and then kept constant for equilibration for 14.5 min. A full MS was acquired in profile mode using a Q Exactive HF-X Hybrid Quadropole-Orbitrap mass spectrometer, operating in the scan range of 375-1400 m/z using 120K resolving power with an AGC of 3x 106 and max IT of 60ms. Data independent analysis followed, using 8 Th windows (39 loop counts) with 15K resolving power with an AGC of 3x 105 and max IT of 22 ms, and a normalized collision energy (NCE) of 26. The mass spectrometry proteomics data have been deposited to the ProteomeXchange Consortium via the PRIDE (21,22) partner repository with the dataset identifier PXD040451.

### MS Data Analysis

Orbitrap raw data was analyzed in DIA-NN 1.8 (Data-Independent Acquisition by Neural Networks) by searching against the reviewed Human Uniprot database (retrieved 4/21) in the library free mode of the software, allowing up to two tryptic missed cleavages. Human Uniprot Database includes 27246 proteins and 21442 genes with 10241864 precursors generated. A spectral library was created from the DIA runs and used to reanalyze them. Parameters regarding peak generation and analysis are defined in the DIA-NN algorithm. DIA-NN default settings have been used with oxidation of methionine residues and acetylation of the protein N-termini set as variable modifications and carbamidomethylation of cysteine residues as fixed modification. N-terminal methionine excision was also enabled. A maximum number of variable modifications set to 3. Both ends are fully tryptic, allowing up to two tryptic missed cleavages. The match between runs feature was used for all analyses and the output (precursor) was filtered at 0.01 FDR. Retention time alignment is performed in DIA-NN. Correction for the mass accuracy is performed for each sample in DIA-NN automatically. Filtering of the quality is based on the false discovery rate of 0.01 at peptide and protein levels. Finally, the protein inference was performed on the level of genes using only proteotypic peptides. The analysis was set for at least one unique peptide per protein. The generated results were processed statistically and visualized in the Perseus software v1.6.15.0 (Max Planck Institute of Biochemistry) and GraphPad Prism 8.0.1. Raw data were filtered based on a minimum of 50% valid values in at least one group of responder/non-responder and log2 transformed. After initial processing non-human genes were excluded from further consideration while missing values were replaced with imputed values which correspond to the limit of detection (LOD). The modified Student’s t-test known as Welch’s Test for Unequal Variances is used. In general, for samples with unequal variance, the adjusted degrees of freedom tend to increase the test power. Differentially expressed proteins (DEPs) were classified as proteins with *p<0.05* (Unequal Welch t-test with S0 cut off 0.1). Proteins with a Welch t-difference above 0 were classified as overexpressed in responders compared to non-responders while proteins with a Welch t-test difference lower than 0 were classified as overexpressed in non-responders compared to responders. Visualization of the obtained results was performed using a Volcano plot.

### Pathway enrichment analysis

To understand the mechanism of response to treatment, pathway enrichment analysis was performed on DEPs between responders and non-responders using Metascape software (MSBio v3.5.20220422). Enrichment analysis parameters were set on a minimum of three genes overlapping between pathways and the input lists. Kyoto Encyclopedia of Genes and Genomes (KEGG) and Reactome and Hallmark (MSigDB) ontologies were used for correlation. Only statistically significant pathways (p-value ≤ 0.05 and minimum enrichment score above 1.5) were taken into account. The obtained results were considered and represented based on biological relevance with respect to RC biology. As a result, the leading term from each group is provided for simplification.

### STRING in silico analysis

The STRING analysis network of DEPs overrepresented in responder/non-responder group was built based on the highest confidence (0.9) evidence from experimental interaction data, co-expression data, gene fusions, gene co-occurrence, gene neighborhood, predictive and knowledge text mining. For easier data processing of 700 DEPs in non-responder group, disconnected nodes in the network were not presented. The analysis was performed using STRING v.11.5 and corresponding images and data downloaded in the original form with statistical significance set at p < 0.05 (23).

### Shortlisting of potential biomarkers

Shortlisting of potential biomarkers was performed using the ROCplotter (www.rocplot.org), an online tool that uses the transcriptome data of a large set of rectal cancer patients (N=284) to find gene expression-based predictive biomarkers. A single database was created by combining published gene expression data from accessible datasets with treatment information (24) Receiver operating characteristic (ROC) curve analysis was performed to assess the predictive accuracy of each gene (25). The observed cohort included 42 patients treated with 5-fluorouracil and radiotherapy and was categorized into responders (N=23) and non-responders (N=19) according to the Response Evaluation Criteria in Solid Tumors (RECIST) criteria. Using a score method devised to assess each probe set for specificity, coverage, and degradation resistance, the optimal microarray probe set to represent a gene was chosen using the JetSet tool (26). ROC curve with p-value<0.05 was considered to evaluate the prediction ability of genes that showed a significant difference between the two groups.

## Results

### Proteomic comparison of Responders and Non-responders

The use of DIA-MS allowed the identification and quantification of more than 3,000 proteins per sample (Supp. Table 1), a significant increase when compared to the 1,000 proteins identified by DDA-MS in rectal cancer FFPE samples (27). In total, 4,269 proteins were identified in 20 rectal cancer FFPE samples (Supp. Table 2). Principal Component Analysis (PCA) indicated that responders had significantly different proteomics profiles than non-responders, as shown in Figure 1a. For comparison, a volcano plot [–log10(P-value) vs. Welch t-test difference] was created to graphically show the proteome changes between the two groups of samples (Figure 1b).

**Figure 1.**
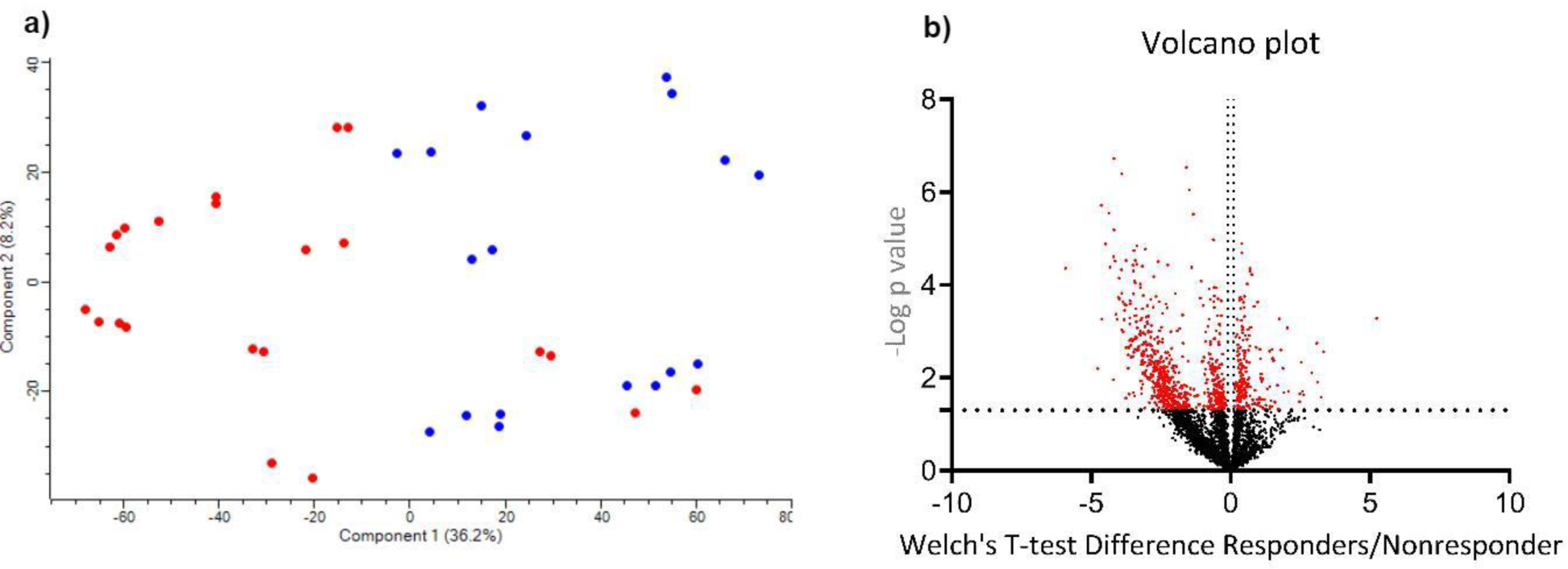
Principal Component Analysis (a) and a Volcano plot (b) of differentially abundant proteins in terms of response to treatment. PCA (a) indicated that responders (blue) compared to non-responders (red) had significantly different proteomics profiles. Proteomics profiles of responders vs. non-responders were compared and results were represented by Volcano plot as - log10 (Welch t-test p-value) plotted against Difference (Responders-Non-responders). Genes upregulated in responders were colored blue, and those upregulated in non-responders were colored red.

Statistical analysis indicated 915 DEPs with statistical significance (p<0.05; S0=0.1) between responders and non-responders. When two groups were compared, DEPs included 700 proteins overexpressed in non-responders and 215 overexpressed in responders (Supp. Table 3; Figure 1b). Tables 2 and 3 exhibit the top 20 examples of DEPs overexpressed in responders (Table 2) and non-responders respectively (Table 3). Using a more stringent statistical setting (p<0.01; S0=0.1), 384 DEPs were found between responders and non-responders, 81 of which were upregulated in responders compared to non-responders, and 303 proteins vice versa (Supp. Table 4).

**Table 2.**
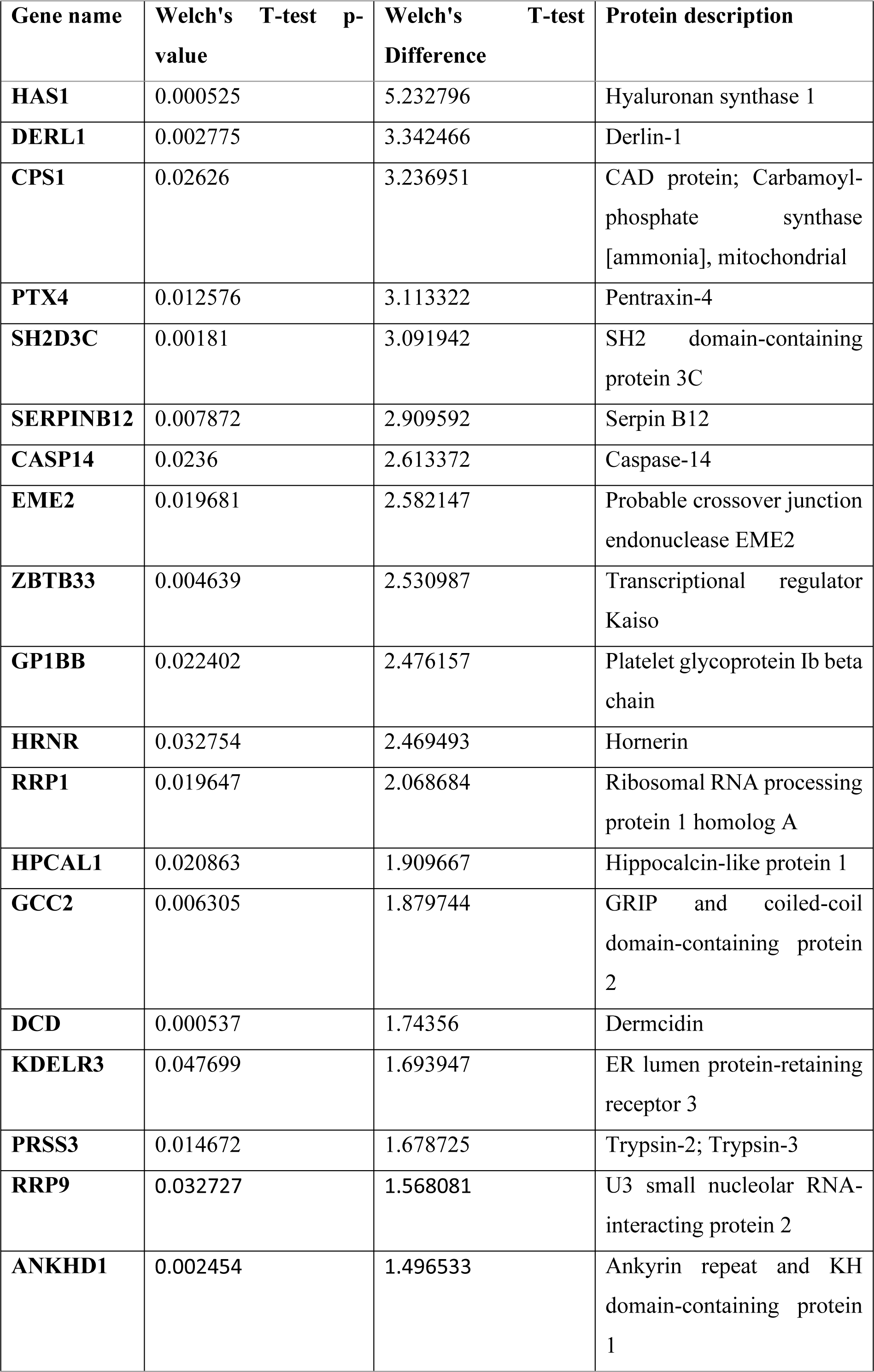
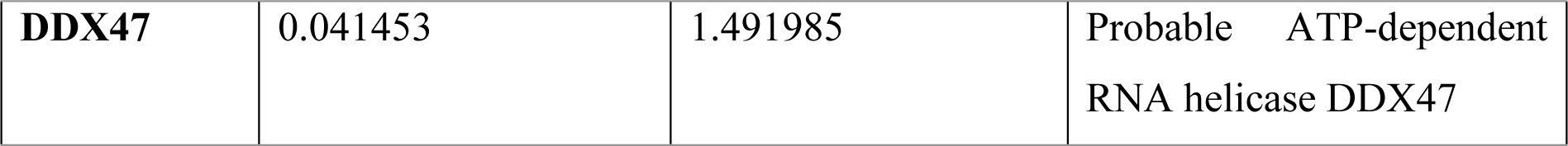
Top 20 DEPs overexpressed in responders compared to non-responders with Welch t-test p-value<0.05.

**Table 3.** Top 20 DEPs overexpressed in non-responders compared to responders with Welch t-test p-value<0.05.

| **Genes name** | **Welch's T-test p-value** | **Welch's T-test Difference** | **Protein description** |
| --- | --- | --- | --- |
| **QPRT** | 4.43E-05 | -5.92343 | Nicotinate-nucleotide pyrophosphorylase [carboxylating] |
| **RBP3** | 0.006363 | -4.76529 | Retinol-binding protein 3 |
| **SPTB** | 1.94E-06 | -4.63391 | Spectrin beta chain, erythrocytic |
| **MOCS2** | 0.00055 | -4.6302 | Molybdopterin synthase sulfur carrier subunit |
| **NHLRC3** | 1.31E-05 | -4.48677 | NHL repeat-containing protein 3 |
| **SMPDL3A** | 2.87E-06 | -4.36289 | Acid sphingomyelinase-like phosphodiesterase 3a |
| **SPTA1** | 4.14E-05 | -4.32445 | Spectrin alpha chain, erythrocytic 1 |
| **CLCA4** | 0.011094 | -4.21054 | Calcium-activated chloride channel regulator 4 |
| **COPS7A** | 2.48E-05 | -4.20115 | COP9 signalosome complex subunit 7a |
| **SYNJ2** | 6.54E-06 | -4.1862 | Synaptojanin-2 |
| **PCTP** | 1.90E-07 | -4.18249 | Phosphatidylcholine transfer protein |
| **GALNT5** | 3.08E-05 | -4.14323 | Polypeptide N-acetylgalactosaminyltransferase 5 |
| **MUC12** | 0.000429 | -4.10598 | Mucin-12 |
| **ACP5** | 0.000559 | -4.07034 | Tartrate-resistant acid phosphatase type 5 |
| **ATG4B** | 0.000189 | -4.01584 | Isoform 2 of Cysteine protease ATG4B |
| **CDHR5** | 7.13E-05 | -4.00168 | Cadherin-related family member 5 |
| **PTGES** | 0.000208 | -3.98789 | Prostaglandin E synthase |
| **TMEM223** | 4.70E-05 | -3.91513 | Transmembrane protein 223 |
| **SORBS3** | 0.000934 | -3.91073 | Vinexin |
| **PLCD1** | 0.000152 | -3.90542 | 1-phosphatidylinositol 4,5-bisphosphate phosphodiesterase delta-1 |

### Pathway enrichment analysis

Enrichment pathway analysis was performed on proteins that were significantly differentially expressed (p<0.05; S0=0.1). Initially, all proteins were chosen regardless of grouping with the goal of better understanding the signaling pathways involved in treatment response (Figure 2a). Further enrichment analysis was carried out on two groups (responders and non-responders) independently in an attempt to explain discrepancies in treatment response. The findings were analyzed and represented based on their biological significance to RC biology. To keep the analysis output concise, only the leading terms of each pathway are shown. Results indicated that some of the leading signaling pathways that correlate with response to nCRT in patients with LARC include the metabolism of RNA, MYC targets, neutrophil degranulation, cellular transport, and response to stimuli. The responder group was characterized by signaling pathways related to cell cycle signaling (metabolism of RNA, synthesis of DNA, DNA strand elongation, mitochondrial translation initiation, chromosome maintenance) as well as MYC targets scores, regulation of expression of SLITs and ROBOs, mTOR1 signaling pathway and unfolded protein response (Figure 2b). The non-responder group was characterized by signaling pathways related to vesicle-mediated transport, neutrophil degranulation, hemostasis, coagulation, heme metabolism, post-translational modifications as well as the metabolism of vitamins, cofactors, and lipids. Signaling pathways related to epithelial-mesenchymal transition and hypoxia which have been associated with an increased risk of metastasis were also found to be important in non-responders (Figure 2c).

**Figure 2.**
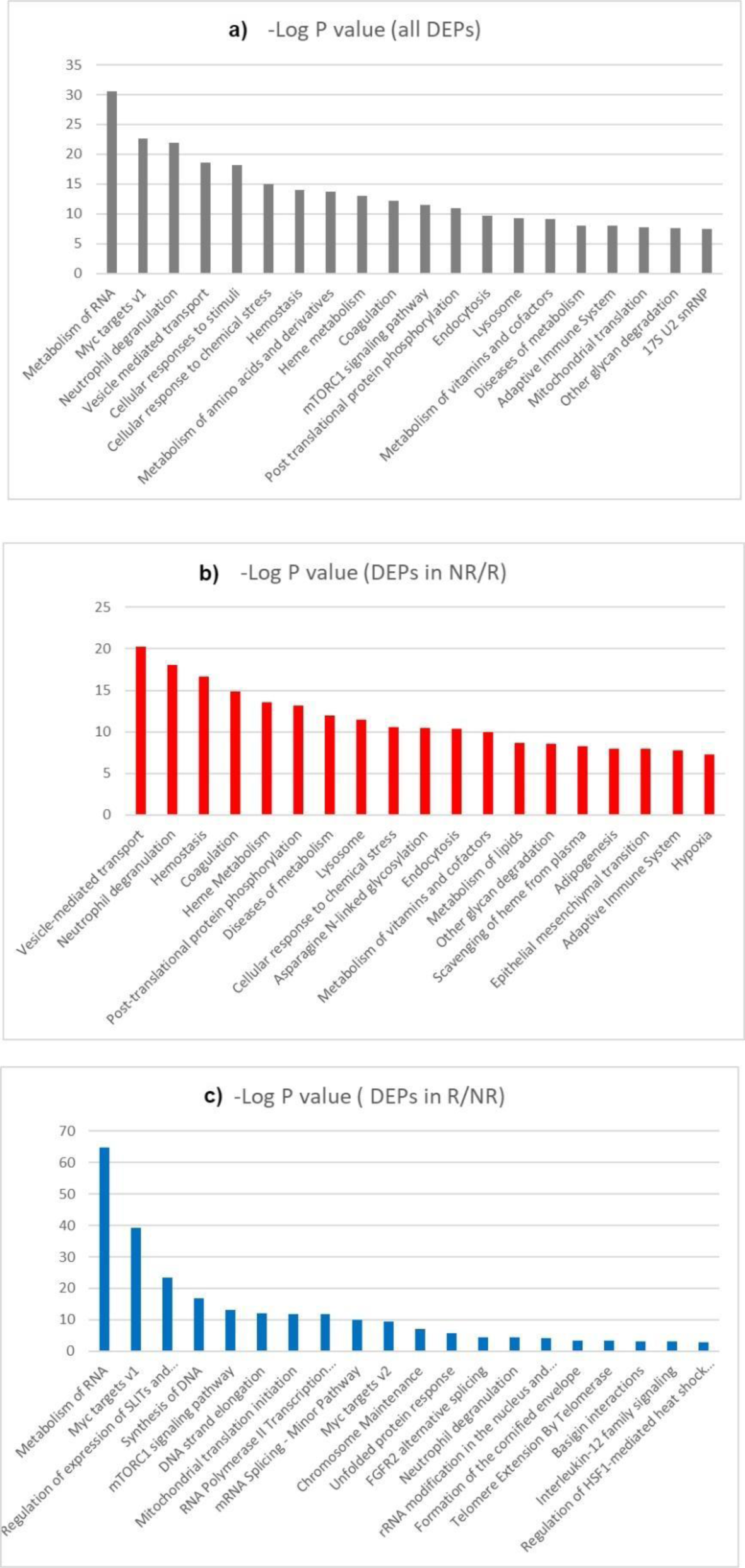
Enrichment pathway analysis of the differentially expressed proteins between responders and non-responders. Only the leading terms from each pathway group are shown, with the size of the bars indicating their importance. The obtained results were considered and represented based on biological relevance with respect to rectal cancer biology. KEGG, Reactome, Corum, and Hallmark databases were used for pathways enrichment analysis. a) signaling pathways characterized by all DEPs; b) signaling pathways altered in responders; c) signaling pathways altered in non-responders.

### STRING in silico analysis

Data obtained reveal several protein-rich groups with several members having high levels of interactions in the responder group (PPI enrichment p-value: < 1.0e-16) indicating that proteins interact with one another more frequently than would be predicted by a randomly selected group of proteins from the genome with the same size and degree distribution (23). We can conclude that there is a strong interaction between proteins involved in information RNA processing and genes whose protein products participate in transcription, especially when it comes to pre-mRNA processing (Supp. Figure 1a) and factors involved in the process of alternative splicing (Supp. Figure 1b). A high degree of interaction is also associated with proteins that participate in the formation of snRNA molecules (Supp. Figure 1b). Another group of proteins that are clearly distinguished based on STRING analysis are the ribosomal proteins of the RPL family (Supp. Figure 1e) and MRPL family (Supp. Figure 1f), as well as PA2G4, which represents the proliferation factor (Supp. Figure 1e). All the mentioned groups of proteins are characterized by a high mutual connection. Another group of proteins includes factors involved in the DNA replication process as well as factors for the organization of the proteasomal system (Supp. Figure 1d and 1c). On the other hand, the group of DEPs overrepresented in the non-responder group is characterized by a much larger number of proteins that are less closely related to each other and the groups cannot be clearly observed. One of the more noticeable groups as central proteins has KRAS, which is related to the YWHA protein family, which participates in the regulation of signaling pathways, as well as RAC1 and MAPK1, which represent one of the main members highly related to other proteins of the group (Supp. Figure 2a). Within the second group (Supp. Figure 2b), proteins involved in fat and lipid metabolism as well as complement components (Supp. Figure 2c) stand out.

### Shortlisting of potential biomarkers based on transcriptomics data

The proteomics results obtained were further examined for discovering promising predictive biomarkers of response to neoadjuvant chemoradiotherapy in patients with LARC. The differential expression of proteins identified in our study was confirmed in transcriptomics datasets. The ROC curve was considered to assess the performance of predictive biomarkers for response to chemoradiotherapy. For this purpose, DEPs obtained after DIA-MS/MS were analyzed using ROCplotter software, and genes with AUC> 0.7, ROC p-value <0.05 and Mann Whitney p-value <0.05 were categorized as promising biomarkers. Out of a total of 915 DEPs, 23 genes met all three criteria. The responder group had the following proteins whose expression was confirmed also at the mRNA level: CRKL, LAP3, THTPA, PES1, PPP2R5E, IFI30, C17orf75, QDPR, RRM2B, USO1, GLRX ARAF, CTBS and SNRPD3. Moreover, the non-responder group had the following proteins COPB1, MGLL, HAS1, TALDO1, DNAH9, KDELR3, HLA-DPB1, RBP3 and STAP2, as presented in Table 4 and Table 5. ROC curves and their discriminatory potential for the mentioned genes are shown in Figure 3 and Figure 4.

**Figure 3.**
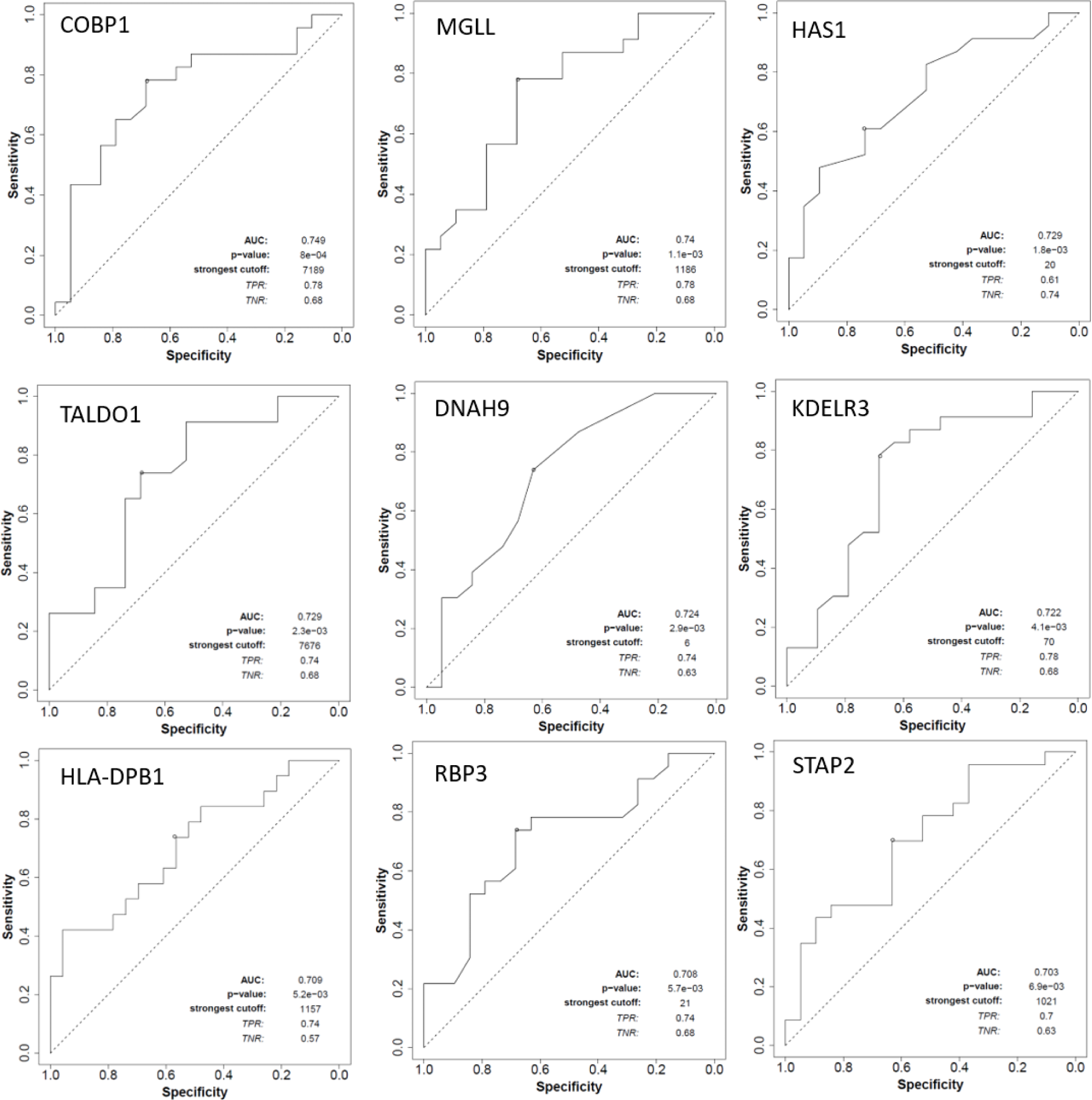
ROC plots of differentially expressed genes overexpressed in non-responder group. ROC plots of genes overexpressed in the non-responder group that met all three criteria and characterized as promising biomarkers on gene expression level AUC> 0.7, ROC p-value <0.05 and Mann Whitney p-value <0.05.

**Figure 4.**
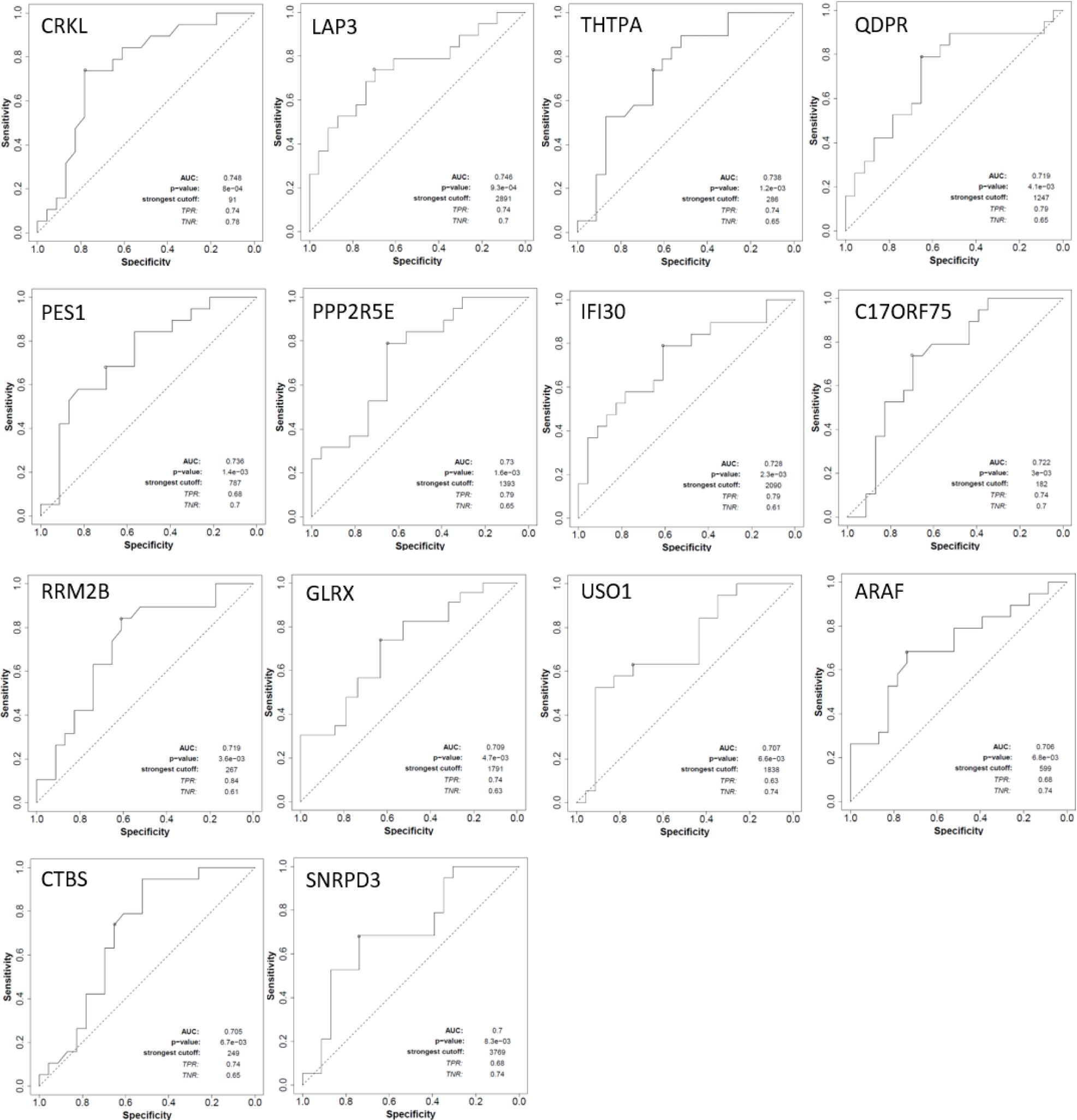
ROC plots of differentially expressed genes overexpressed in the responder group. ROC plots of genes overexpressed in the responder group that met all three criteria and characterized as promising biomarkers on gene expression level AUC> 0.7, ROC p-value <0.05, and Mann Whitney p-value <0.05.

**Table 4.** Shortlisted DEPs with characteristics of promising biomarkers enriched in the responder group compared to the non-responder group. Genes with a ROC p-value less than 0.05, an AUC greater than 0.7, and a Mann-Whitney p-value less than 0.05 were characterized as promising biomarkers.

| **Genes** | **FOLD change** | **Mann Whitney test p-value** | **AUC** | **ROC p-value** |
| --- | --- | --- | --- | --- |
| **CRKL** | 1.4 | 0.0063 | 0.748 | 8.00E-04 |
| **LAP3** | 1.5 | 0.0059 | 0.746 | 9.30E-04 |
| **THTPA** | 1.4 | 0.0089 | 0.738 | 1.20E-03 |
| **PES1** | 1.2 | 0.0096 | 0.736 | 1.40E-03 |
| **PPP2R5E** | 1.2 | 0.01 | 0.73 | 1.60E-03 |
| **IFI30** | 1.5 | 0.011 | 0.728 | 2.30E-03 |
| **C17orf75** | 1.3 | 0.015 | 0.722 | 3.00E-03 |
| **QDPR** | 1.3 | 0.013 | 0.719 | 4.10E-03 |
| **RRM2B** | 1.3 | 0.016 | 0.719 | 3.60E-03 |
| **GLRX** | 1.7 | 0.02 | 0.709 | 4.70E-03 |
| **USO1** | 1.2 | 0.022 | 0.707 | 6.60E-03 |
| **ARAF** | 1.2 | 0.024 | 0.706 | 6.80E-03 |
| **CTBS** | 1.3 | 0.024 | 0.705 | 6.70E-03 |
| **SNRPD3** | 1.2 | 0.027 | 0.7 | 8.30E-03 |

**Table 5.** Shortlisted DEPs with characteristics of promising biomarkers enriched in the non-responder group compared to the responder group. Genes with a ROC p-value less than 0.05, an AUC greater than 0.7, and a Mann-Whitney p-value less than 0.05 were characterized as promising biomarkers.

| **Genes** | **FOLD change** | **Mann Whitney test p-value** | **AUC** | **ROC p-value** |
| --- | --- | --- | --- | --- |
| **COPB1** | 1.1 | 0.0061 | 0.749 | 4.00E-04 |
| **MGLL** | 1.3 | 0.0083 | 0.74 | 1.10E-03 |
| **HAS1** | 1.9 | 0.012 | 0.729 | 1.80E-03 |
| **TALDO1** | 1.2 | 0.012 | 0.729 | 2.30E-03 |
| **DNAH9** | 1.4 | 0.013 | 0.724 | 2.90E-03 |
| **KDELR3** | 1.6 | 0.015 | 0.722 | 4.10E-03 |
| **HLA-DPB1** | 1.8 | 0.02 | 0.709 | 5.20E-03 |
| **RBP3** | 1.8 | 0.022 | 0.708 | 5.30E-03 |
| **STAP2** | 1.2 | 0.025 | 0.703 | 6.90E-03 |

## Discussion

Understanding the molecular features associated with response to neoadjuvant chemoradiotherapy is an unmet clinical need in LARC. The DIA-MS offers unprecedented proteome coverage for FFPE samples. In comparison to traditional data-dependent acquisition mass spectrometry (DDA-MS), DIA-MS enabled in-depth characterization and higher reproducibility in one injection (28). As a main difference, in DIA, the mass spectrometer is set up to cycle through a predefined set of precursor isolation windows, consistently fragmenting all the precursors within the target mass range, compared to choosing the most abundant precursor ions in the data-dependent manner (29,30). DIA-MS also enables the in-depth study of the proteome from FFPE tissue samples, which represented a major challenge because of damage caused by the fixation protocol (31). By exploring the dynamic phenotypic characteristics of tumor cells before therapy and tumor response to therapy, DIA-MS allows us to characterize response mechanisms and thus enable patient monitoring and more effective treatment. Using DDA-MS, the identification of low-abundance proteins, that may be crucial for altering the activity of signaling pathways and may cause differences in response to therapy, is a challenging task. FFPE samples are routinely used for DDA-MS analysis, and the use of FFPE samples for the DIA-MS method is increasing (32). In contrast to fresh frozen (FF) tissue, FFPE samples undergo protein cross-linking during standard preservation protocol, and due to that is challenging to analyze native proteins. For this reason, it is necessary to take care to reverse cross-linked proteins, which is part of the standard protocol used in this study (33). Comparing the results obtained when it comes to FF tissue versus FFPE samples indicates a high correlation between the results, which makes FFPE samples a good alternative to FF samples (34,35). On the other side, FFPE samples are more suitable for retrospective studies and are easily accessed during everyday clinical routines (32).

Several studies have been published on the proteomic profile of locally advanced rectal cancer cells in terms of response to therapy (27,36,37). Due to technical limitations, results obtained in the previous studies showed a restricted number of identified proteins, while DIA-MS/MS offered an in-depth characterization of rectal cancer tissue enabling molecular characterization and profiling the response. During the last decades, 2D DIGE electrophoresis was the method used for proteomic analysis of the cells (36), which was followed by DDA-MS which showed a significant improvement compared to 2D DIGE. With method improvement, we were able to identify 3 to 4 times more DEPs. DIA-MS/MS showed undoubtedly in-depth characterization of the proteomic profile. Our study included analysis of discrete and well-characterized clinical samples of rectal cancer in order to identify the maximum number of different molecular features potentially associated with response. Thus, a significantly higher number of DEPs was identified compared with a previous DIA-MS/MS proteomics study performed in a rectal cancer patients cohort (37).

The use of DIA-MS allowed the identification and quantification of more than 3,000 proteins per sample in general, a significant increase when compared to the 1,000 proteins identified by Data Dependent Acquisition-MS in LARC FFPE samples. In total 4,269 proteins were identified in 20 rectal cancer FFPE samples. Principal Component Analysis (PCA) indicated that responders had a significantly different proteomics profile than non-responders. Statistical analyses comparing the two groups resulted in the identification of 915 differentially expressed proteins (215 in responders and 700 in non-responders) (p<0.05).

Some of the top 20 proteins that are differentially overexpressed in the group of non-responders versus responders are drug targets used in the treatment of some other pathological conditions. Niacin, for instance, is used to treat cholesterol and its target is the QPRT protein. Talniflumate, an anti-inflammatory drug, is used to treat cystic fibrosis and its target is CLCA4. Alteplase and filgrastim, which target GALNT5, are used as a thrombolytic agent, an agent for preventing infections and increasing the level of neutrophils, respectively. Esomeprazole and Nimodipine whose target is ATG4B and are used in treating stomach acid secretions and symptoms resulting from a ruptured blood vessel in the brain. Aspirin, NSAIDs, and COX2 inhibitors are used in the treatment of lowering the temperature and their target is PTGS2 (38,39). The question of the discriminatory effect of gene expression is raised, as well as whether the use of these drugs can be used as a part of initial treatment and would lead to a better response to therapy by treating patients with locally advanced rectal cancer together with standard neoadjuvant chemoradiotherapy.

The therapy approach used for LARC includes radiotherapy in combination with 5-fluorouracil-based chemotherapy followed by surgery. The effect of radiotherapy is based on the formation and reaction of molecular fragments such as free radicals and excited molecules, which further leads to damage to DNA molecules. It is considered that the cell is most sensitive to radiation during mitosis and in the early G1 phase, while it is most resistant in the S phase of the cell cycle. The above-mentioned effects are first reflected in the cell cycle and the overcoming of toxic effects is reflected through the reparation of the resulting damage. 5-fluorouracil as a standard chemotherapeutic drug is an antimetabolite that is metabolized by the liver. 5-fluorouracil enters the cell via a transporter human nucleoside transporter (hENT1, hENT2), human organic anion transporter 2 (hOAT2), and myeloid-related proteins through efflux of 5-FdUMP (MRP-5 and MRP-8) as well as through paracellular transport (40). 5-FU has three active metabolites (5-FdUMP, 5-FdUTP, and 5-FUTP) that result from the metabolic conversion of 5-FU. 5-FdUMP and 5-FdUTP act at the level of DNA, causing damage to DNA molecules, while 5-FUTP integrates into RNA and exerts an antiproliferative effect. Our finding confirmed the importance of DNA metabolism and related signaling pathways which were altered in both responder and non-responder groups in terms of response to therapy. It is shown that signaling pathways related to DNA strand elongation and synthesis of DNA are affected in patients with poor response to treatment. It has been reported in mice treated with 5-FU, that there is a dependence on the cytotoxic effect of nucleoside balance. Namely, the administration of 5-FU with uridine led to an inhibition of the cytotoxic effect compared to the administration of thymidine. This fact highlighted that 5-FU affects RNA and mRNA splicing as well (41,42). Our results suggested that the mRNA splicing-minor pathway, as well as rRNA modification in the nucleus and cytosol, and the metabolism of RNA pathways, are affected in good responders to nCRT. A novel stress response mechanism called decreased splicing efficiency and global intron retention may help malignant cells survive treatment under normal homeostatic conditions. Data obtained in this study indicated that cellular response to chemical stress correlated with a lack of response to therapy. Numerous investigations have shown that spliceosome proteins can be produced in extracellular space when chemotherapy and radiation are employed (43,44). Increased activity of mRNA splicing in the responder group can neutralize the negative effect of 5-FU which can potentially lead to a good response to treatment. Pre-mRNA alternations might induce altered gene expression, pre-mRNA splicing dysregulation, protein post-translational modification, and secretion (45). According to our data, patients with altered signaling pathways related to post-translational protein phosphorylation are suggested to have significance for poor response, while mRNA splicing, RNA Polymerase II Transcription Termination, and metabolism of RNA were pathways enriched in the responder group.

In all eukaryotes, the spliceosome, a cellular device that removes introns from precursor mRNAs (pre-mRNAs), contains the U2 small nuclear ribonucleoprotein (snRNP). The most often altered splicing component in malignancies, U2, is incredibly dynamic. It has five non-coding snRNAs (U1, U2, U4, U5, and U6) that use base pairing to identify intronic splice sites, coordinate the assembly of protein splicing components, and catalyze the cleavage and ligation events (46). It has been shown that during treatment, 5-FU can be incorporated in spliceosome U2 snRNA at pseudouridylated sites to inhibit U2 pseudouridilation and subsequent pre-mRNA splicing (47). Incorporation of 5-FU into U2 snRNA blocks pseudouridylation and pre-mRNA splicing in vivo was obtained in our study suggesting that metabolism of 17s U2 snRNP is significant for the response to treatment to nCRT in LARC.

Signaling pathways related to Myc targets have also been found to be significant for the good response to nCRT in our LARC cohort. The cell cycle, proliferation, differentiation, apoptosis, and miRNA activation are all crucially regulated by the Myc oncogene, a transcription factor that supports that cell cycle-related signaling pathways play a crucial role in response to treatment (48). Signaling pathways which include chromosome maintenance and telomere extension by telomerase were found to be affected in good response to treatment which is in correlation with previous findings. Additionally, telomere shortening is accelerated by oxidative stress and accumulated reactive oxygen species (ROS) because SSB in telomeres cannot be repaired as effectively (49). This observation suggested that changes at the chromosome level can lead to a good response to treatment and increase the efficiency of RT. Additionally, mTOR, a key regulator of cell growth and division in healthy conditions can be inappropriately activated in tumor cells and thus promote tumor cell growth, metastasis, and invasion of fresh healthy tissues (50). Our findings showed that the signaling pathway mTORC1 was altered in the group of good responders. Analysis of DEPs provided a potential scenario that included the downregulation of genes related to the mTORC1 pathway in responders or overexpression of mTORC1-related genes in non-responders. Considering available data, both scenarios will lead to poor response to treatment. It was shown that activation of mTOR signaling pathways correlates with drug resistance in multiple cancer types (51,52).In metabolic homeostasis, protein and lipid synthesis, glycolysis, mitochondrial biogenesis, and lysosome biogenesis, mTORC1 is essential. The translation is directly influenced by the regulation of different transcription factors. Through metabolic pathway proteins, it controls nucleotide synthesis and glucose metabolism. Additionally, it controls both autophagy and proteasome assembly (53). The mTORC2 primarily controls glucose metabolism, apoptosis, cell migration, cytoskeletal reorganization, and cell proliferation (54,55). According to our results, signaling pathways related to protein and lipid synthesis, glycolysis, mitochondrial biogenesis, and lysosome biogenesis correlate with poor response to nCRT which is in compliance with previous findings. An in-depth characterization of mTORC1 signaling pathways is needed to shed light on the exact biochemical mechanism that leads to good/poor responses to treatment.

During the process of immune defense against damage that CRT may induce, cytokines and growth factors are released, which play a significant role in the production of ROS, primarily superoxide, hydrogen peroxide, and nitrogen (II)-oxide (56). Preclinical investigations have shown that cytokines, such as interleukins (IL-2, IL-12) might aid in the radiosensitization of cells and modulation of antitumoral response (57). IL-12 achieves its antitumor activity by promoting the immune response via the activation of natural killer cells, and cytotoxic T-cells and exerting an antiangiogenic effect (45,58). In a study performed by Heeran et al., increased levels of IL-12 were detected in the blood of LARC patients compared to healthy individuals. This suggests the potential for promoting an immune response that may lead to improved treatment outcomes (59,60). Our results indicated that the immune response might play an important role in predicting response to therapy. Signaling pathways associated with IL-12 were found to correlate with a good response to therapy while signaling pathways related to the adaptive immune system were related to a poor response. Heeran et al. highlighted the association of inflammation with obesity status in rectal cancer patients in terms of lowering the level of inflammatory factors released from TME (59). Our data correlated adipogenesis with poor response treatment. As the increased synthesis of adipose tissue is directly correlated with an increase in body weight, it might further lead to a worse outcome of therapy. A study by Lee et al. was shown that obesity represents an independent predictor of cCR, which contradicts our results (61). In some studies, no clear correlation was found between obesity and rectal cancer treatment outcomes (62).

Angiogenesis plays an important role in the development of cancer, and it has been shown that hypoxia is related to poor response to treatment. During hypoxic environment angiogenesis is activated, thus the blood supply to the tumor and the rate of progression increase, which might impair the effect of the therapy. The results obtained in this study indicate that signaling pathways associated with heme uptake from plasma, heme metabolism, and hemostasis are highly associated with poor response to therapy. Due to a change in the metabolism of heme, which plays a role in the transport of oxygen to tissues, tumor progression might be enhanced leading to the lower effectiveness of therapy. We also detected that the response to CRT and coagulation might correlate. Previous findings are in line with our data, indicating that cancer cells activate the coagulation process and that hemostatic factors might play an important role in tumor progression (63).

It has also been shown that vitamins can act as modulators of the immune response and thus participate in the development and progression of cancer (64–66). In our study results, it was shown that in patients with a poor response to therapy, the signaling pathway of vitamin and cofactor metabolism is altered, which reinforces the previously reported data.

Slit-Robo signaling also plays an important role in angiogenesis. Endothelial cells only express the vertebrate Robo4 gene, which has been linked to controlling angiogenesis and blood vessel permeability (67,68). In our cohort, it was shown that the regulation of SLITs and ROBOs is highly correlated with a good response to nCRT. Slit/Robo signaling has both pro- and anti-angiogenic functions, therefore its effect in angiogenesis depends on the environment. It has been shown that Slit2 promotes angiogenesis via the Robo1 receptor, while on the contrary via Robo4 it participates as an inhibitor of endothelial migration, which benefits a good response to therapy. It was also reported that Split3 promotes angiogenesis (69). In terms of good response to treatment and favorable outcome, inhibition of endothelial migration angiogenesis is preferred. In support of those observations, our results indicated the importance of epithelial-mesenchymal transition in patients with a poor response to therapy. Hypoxia has also been linked with neutrophil degranulation. In hypoxic conditions, degranulation occurs, and released factors affect tumor progression(70). According to our results, the neutrophil degranulation pathway was affected in both the responder and non-responder groups which highlights its importance in this process and warrants further functional studies.

In rectal cancer patients receiving neoadjuvant CRT, it was shown that high expression of FGFR2 was associated with an advanced tumor stage, a poor treatment response, and lower survival. (71). The DIA-MS approach indicated the importance of FGFR2 alternative splicing in good response to treatment, which implies that exploring its variants might be useful for the prediction of a good outcome.

The highest correlation with poor response to treatment was shown for signaling pathways related to vesicle-mediated transport, endocytosis, and lysosomes. Proteins and other cargo must be carried through the cell via a cellular transport mechanism in which the transported materials are conveyed in membrane-bound vesicles. The vesicle lumen or the vesicle membrane is where transported compounds are contained (72). Exosomes released from cell membrane has a significant role in tumor proliferation, differentiation, metastasis, and resistance to chemotherapy and radiation by transporting biomolecules (proteins, lipids, deoxyribonucleic acid, and ribonucleic acid) throughout the tumor microenvironment (73). The results obtained in this study indicated a great potential for exploring intercellular communication in the tumor microenvironment as well as in the tumor when profiling response to therapy. The synergistic effect of these inter-relations has not yet been clarified and would also be validated in future functional studies by our group.

Literature data from the past several decades identified altered glycosylation as a sign of malignancy. It was shown that glycosylation acts as a mediator of the inflammatory response (74). Correlation between glycosylation and radioresistance was already shown in laryngeal carcinoma and gliomas (75). According to our results, glycan degradation and asparagine N-linked glycosylation might play an important role in the poor response of LARC patients to nCRT.

Linking the transcriptomic and proteomic profiles of the cell is an important parameter for understanding the molecular basis of the response to therapies. The software ROC plotter showed a low correlation between gene expression and the proteome profile of the tested samples. Out of 915 differentially expressed proteins, only 23 showed promising discriminatory potential when it comes to gene expression. It should be kept in mind that the transcriptomics data were obtained from samples of different demographics and ethnic origin, therefore further validation on patient samples within our cohort should be performed. The translation of research from protein to gene expression and confirmation of the obtained candidates would enable a better cost-effectiveness approach and thus a more efficient selection of patients when it comes to predicting the response to neoadjuvant chemoradiotherapy. Gene expression analysis shows high sensitivity when it comes to FFPE tissue analysis and would be more likely to be performed during everyday practice. At the same time, further analysis of the gene expression profile and correlation with a proteomic profile of the tissue a would enable more detailed investigation of the mechanism behind response treatment which is still unclear.

Based on all obtained results, we conclude that there is a statistically significant difference between the proteomic profiles of LARC patients who respond well and poorly to nCRT. As all patients require surgery after nCRT per current guidelines, profiling of adequate biomarkers of response is a pressing matter. Further validation of target signaling pathways detected in this study that might have an effect on the response to nCRT is planned on a prospective cohort of LARC patients at the Institute for Oncology and Radiology of Serbia, to ensure more efficient and cost-effective treatment of patients, while maximizing their quality of life.

## Conclusions

Data independent acquisition mass spectrometry (DIA-MS) analysis of FFPE LARC samples offered unprecedented in-depth proteomics characterization and enabled the identification of new molecular features associated with response to nCRT. The observed differentially expressed proteins and biological processes constitute findings with a high potential for future improvement of LARC patient management.

## Data Availability

All non-patient sensitive data produced in the present work are contained in the manuscript. The mass spectrometry proteomics data have been deposited to the ProteomeXchange Consortium via the PRIDE partner repository with the dataset identifier PXD040451.

https://www.ebi.ac.uk/pride/archive

## Acknowledgments

This study was funded by the Horizon Europe STEPUPIORS Project (HORIZON-WIDERA-2021-ACCESS-03, European Commission, Agreement No. 101079217) and the Ministry of Education and Science of the Republic of Serbia (Agreement No. 451-03-47/2023-01/200043). This article is based upon work from COST Action CA17118, supported by COST (European Cooperation in Science and Technology); www.cost.eu. We acknowledge the support of this work by the project “The Greek Research Infrastructure for Personalised Medicine (pMED-GR)” (MIS 5002802) which is implemented under the Action “Reinforcement of the Research and Innovation Infrastructure”, funded by the Operational Programme “Competitiveness, Entrepreneurship, and Innovation” (NSRF 2014-2020) and co-financed by Greece and the European Union (European Regional Development Fund). The mass spectrometry proteomics data have been deposited to the ProteomeXchange Consortium via the PRIDE (15–17) partner repository with the dataset identifier PXD040451.

## Study approval

This study has been approved by the Ethics Committee of the Institute for Oncology and Radiology of Serbia and all patients signed informed consent. All experiments have been performed in accordance with the Helsinki Declaration of 1975, as revised in 2013.

---

**Supp. Material 1.** Sample ID with corresponding category (responder/non-responder) and SDS page after protein separation

**Supp. Table 1. Identification and quantification of proteins by employing the DIA-MS approach.**

**Supp. Table 2.** Identification and quantification of proteins with statistics.

**Supp. Table 3.** Identification and quantification of 915 DEPs with statistical significance (p<0.05; S0=0.1) between responders and non-responders.

**Supp. Table 4.** Identification and quantification of 384 DEPs with more stringent statistical significance (p<0.01; S0=0.1) between responders and non-responders.

**Supplementary Figure 1.**
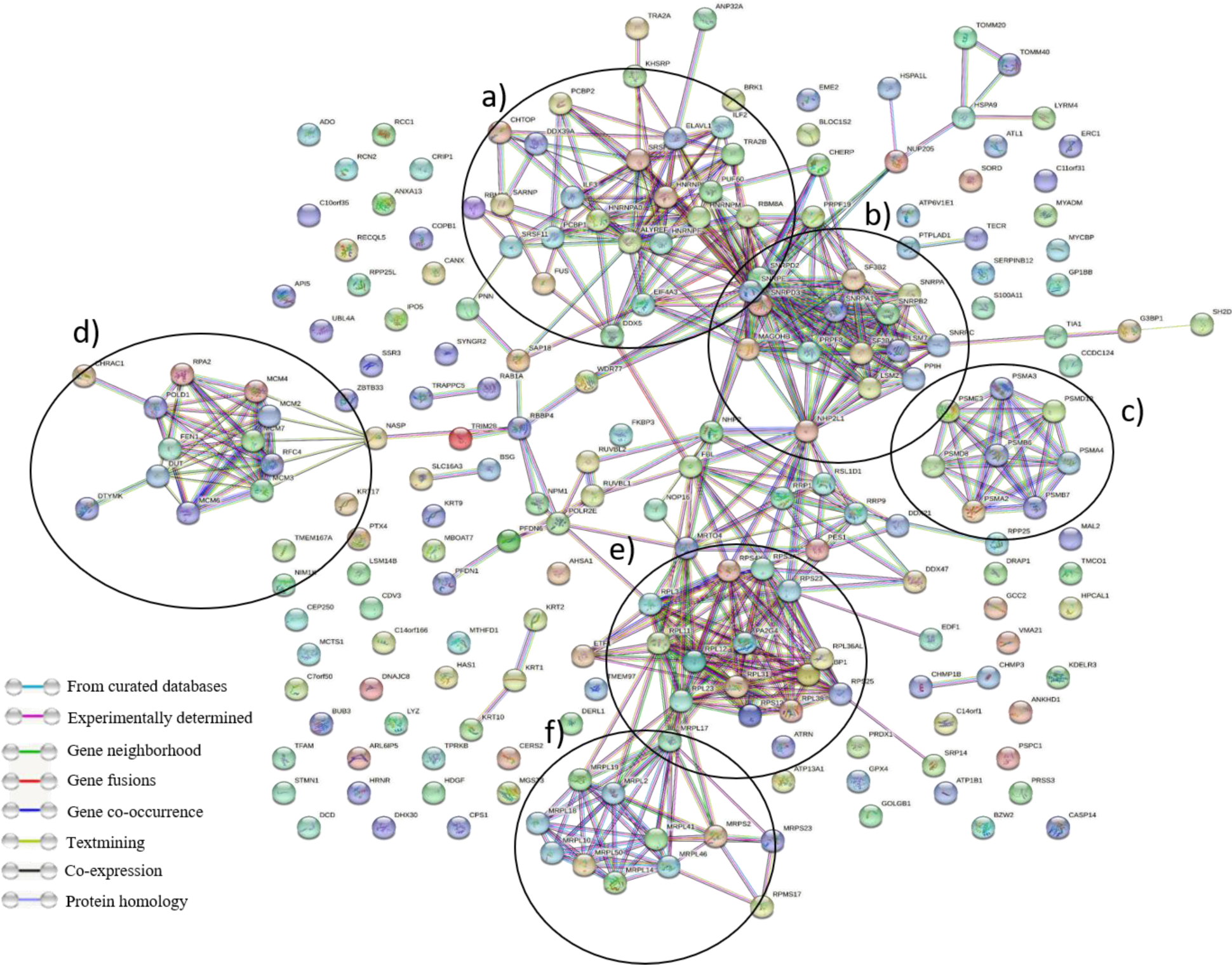
Representation of DEPs overrepresented in responder compared to non-responder group by STRING software.

**Supplementary Figure 2.**
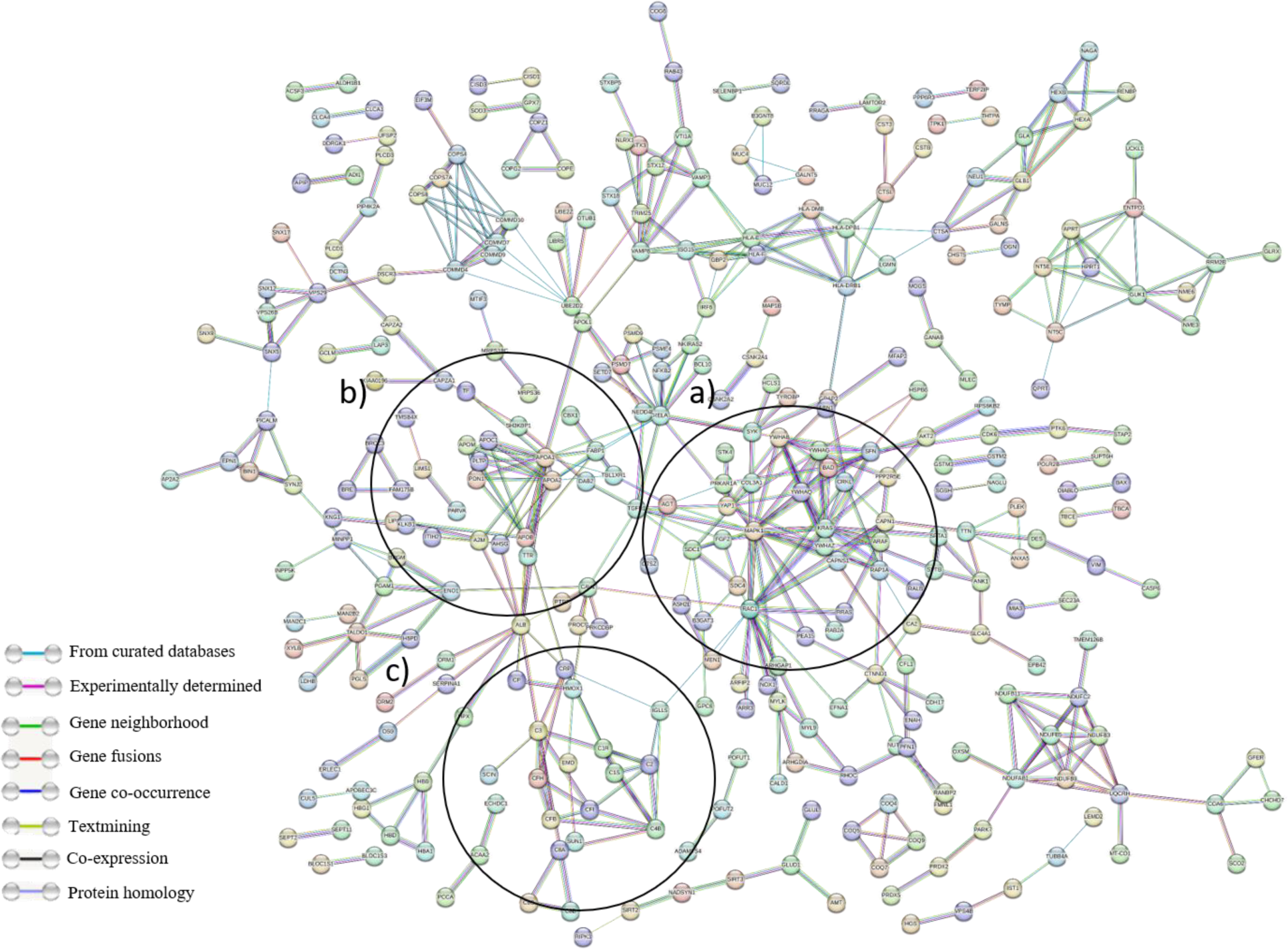
Representation of DEPs overrepresented in non-responder compared to responder group by STRING software.

